# Shotgun mass spectrometry-based lipid profiling identifies and distinguishes between chronic inflammatory diseases

**DOI:** 10.1101/2021.03.08.21252659

**Authors:** Rune Matthiesen, Chris Lauber, Julio L. Sampaio, Neuza Domingues, Liliana Alves, Mathias J. Gerl, Manuel S. Almeida, Gustavo Rodrigues, Pedro Araújo Gonçalves, Jorge Ferreira, Cláudia Borbinha, João Pedro Marto, Marisa Neves, Frederico Batista, Miguel Viana-Baptista, Jose Alves, Kai Simons, Winchil L. C. Vaz, Otilia V. Vieira

**Affiliations:** iNOVA4Health, CEDOC, NOVA Medical School, NMS, Universidade Nova de Lisboa, 1169-056 Lisboa, Portugal; Lipotype GmbH, Tatzberg 47, 01307, Dresden, Germany; Hospital Santa Cruz, Centro Hospitalar de Lisboa Ocidental, Av. Prof. Dr. Reinaldo dos Santos, 2790-134, Carnaxide, Portugal; Department of Neurology, Hospital de Egas Moniz, Centro Hospitalar de Lisboa Ocidental, Rua da Junqueira 126, 1349-019, Lisboa, Portugal; Hospital Dr. Fernando da Fonseca, IC 19, 2720-276, Amadora, Portugal

**Keywords:** cardiovascular disease, ischemic stroke, systemic lupus erythematosus, dyslipidemia, lipid profiling, lipid biomarker

## Abstract

**Background:** Inflammation impacts several acute and chronic diseases causing localized stress and cell death, releasing tissue-specific lipids into the circulation from inflamed cells and tissues. The plasma lipidome may be expected to reflect the type of inflammation and the specific cells and tissues involved. However, deep lipid profiles of major chronic inflammatory diseases have not been compared.

**Methods:** We compare the plasma lipidomes of patients suffering from two etiologically distinct chronic inflammatory diseases, atherosclerosis-related cardiovascular disease (CVD) including ischemic stroke (IS), and systemic lupus erythematosus (SLE), to each other and to age-matched controls. The controls had never suffered from any of these diseases. Blood plasma lipidomes were screened by a top-down shotgun MS-based analysis without liquid chromatographic separation. Lipid profiling based on MS was performed on a cohort of 427 individuals. The cohort constitutes 85 controls (control), 217 with cardiovascular disease (further classified into CVD 1-5), 21 ischemic stroke patients (IS), and 104 patients suffering from systemic lupus erythematosis (SLE). 596 lipids were profiled which were quality filtered for further evaluation and determination of potential biomarkers. Lipidomes were compared by linear regression and evaluated by machine learning classifiers.

**Results:** Machine learning classifiers based on the plasma lipidomes of patients suffering from CVD and SLE allowed clear distinction of these two chronic inflammatory diseases from each other and from healthy age-matched controls and body mass index (BMI). We demonstrate convincing evidence for the capability of lipidomics to separate the studied chronic and inflammatory diseases from controls based on independent validation test set classification performance (CVD vs control - Sensitivity: 0.90, Specificity: 0.98; IS vs control - Sensitivity: 1.0, Specificity: 1.0; SLE vs control – Sensitivity: 1, Specificity: 0.88) and from each other (SLE vs CVD □ Sensitivity: 0.91, Specificity: 1). Preliminary linear discriminant analysis plots using all data clearly separated the clinical groups from each other and from the controls. In addition, CVD severities, as classified into five clinical groups, were partially separable by linear discriminant analysis. Notably, significantly dysregulated lipids between pathological groups versus control displayed a reverse lipid regulation pattern compared to statin treated controls versus non treated controls.

**Conclusion:** Dysregulation of the plasma lipidome is characteristic of chronic inflammatory diseases. Lipid profiling accurately identifies the diseases and in the case of CVD also identifies sub-classes. Dysregulated lipids are partially but not fully counterbalanced by statin treatment.

## Introduction

Lipids in the blood plasma reflect diet and metabolic characteristics of an individual but are also known to regulate inflammatory responses both positively and negatively. In the case of inflammatory processes cell stress and death also release internal cellular lipids into the blood. It might, therefore, be expected that chronic inflammatory diseases alter the plasma lipidome in a manner that is characteristic of the chemistry of the cells/tissues primarily involved in the disease. With this reasoning we examined the lipidomes of patients suffering from cardiovascular diseases (CVD), including ischemic stroke (IS), and systemic lupus erythematosus (SLE) and compared them with the lipidome of age-matched controls who were not known to suffer, or to have suffered, from any of these diseases.

The role of inflammation and lipids in atherosclerosis is well established [1-3]. There is a reasonably large amount of literature [3-25] on the profiling of lipids in blood plasma and atherosclerotic plaques in CVD cohorts. The two main variables in these studies have been the number of lipids that have been profiled and the size of the cohorts [4, 7]. The emphasis of previous studies has been on the association of plasma lipid species with CVD-risk stratification and CVD-related mortality [5, 6, 8-20], improved classification of stable and unstable CVD states [21], correlation with established diagnostic tools for CVD [18, 22], genetic risk factors for CVD [5, 18, 23], association of CVD with co-morbidities [13, 16, 24, 25], and association of CVD with changes in lipid biochemistry in the blood plasma [26]. In general, the different lipidomic profiling studies differ in the methods and definition of clinical outcome subjected to lipidomic based classification. This means that the clinical outcome and lipids profiled differ across studies. Consequently, consensus targets from multiple studies are still unobtainable. Nevertheless, common to all studies are the highly sensitive changes in lipid profiles depending on the clinical outcome evaluated. Table S1 summarizes individual lipids associated with CVD or CVD risk factors from past studies.

SLE is a chronic inflammatory disease. SLE etiology displays multifactorial characteristics and the molecular mechanisms of this disorder are largely unknown [27-29]. A notable aspect of this disease is the excessive production of reactive oxygen species that oxidize cellular lipids producing derivatives that cause dyslipidemia and dyslipoproteinemia [30, 31]. The dyslipoproteinemia signature has been identified in SLE patients with markedly increased age-specific incidence of cardiovascular disease [31]. Lipidomics of the plasma of SLE patients has been previously reported [32, 33].

In this work we enquired whether it would be possible to characterize the lipidomic profile of different manifestations of essentially the same clinical disease (different degrees of CVD, including IS) and whether the lipidomic profile of two distinct diseases that only had inflammation as a common characteristic could be distinguished from each other and used as a diagnostic identifier. Furthermore, the accuracy as a diagnostic identifier was evaluated on an independent test data set.

Despite the preliminary efforts and promising results based on LC-MS lipid profiling, identifying individuals at risk for stroke and cardiovascular events from a healthy control population remains a challenge. Time invested per sample constitutes a barrier for large scale clinical validation and implementation of lipid profiles (e.g. the LC dimension of previous studies is time intensive). In this study we evaluated classification performance of lipid profiling based on shotgun MS without LC lipid separation prior to MS. A total of 596 lipids were profiled in a total of 427 individuals. Remarkable separations between pathological groups and controls were obtained by partial least square classifiers evaluated on an independent validation data set. In addition, dysregulated lipids in CVD and stroke appear partially opposed by statin treatment.

## Methods

### Patient samples

Plasma samples were obtained from a total of 427 individuals. Baseline characteristics are outlined in Table 1. Control (n = 85) were taken from the population of the Coimbra and Lisbon, Portugal, regions. They satisfied the criterion that they had never had any CVD-or SLE-related health complaints. The CVD patients (n = 238) were divided into 6 groups. CVD1 (n = 61) contains individuals who went to the hospital with chest pain but had no indicators for stable angina pectoris, unstable angina pectoris or myocardial infarction. CVD2 (n = 82) are patients with stable angina pectoris (SAP). CVD1 and CVD2 are defined according to the ACCF/AHA/ACP/AATS/PCNA/SCAI/STS guidelines [34]. CVD3 (n = 20) contains patients with unstable angina pectoris, CVD4 (n = 34) are patients who suffered an acute myocardial infarction with no ST-elevation in ECG, and CVD5 (n = 20) are patients who suffered acute myocardial infarction with ST-elevation in ECG [35, 36]. CVD3, CVD4, and CVD5, together, may be classified as patients with an acute coronary syndrome (ACS). CVD1 through CVD5 groups were all obtained from Hospital Santa Cruz, Carnaxide, Portugal. Acute ischemic stroke (IS) (n = 21) were patients admitted at the emergency room of the Centro Hospitalar de Lisboa Ocidental, Lisbon, Portugal, who suffered from acute ischemic stroke. The SLE cohort (n = 104) were patients from Hospital Dr. Fernando Fonseca, Amadora, Portugal. The inclusion criteria were all patients diagnosed with the pathology and above 18 years old. The exclusion criteria were the existence of serious renal and hepatic pathologies, cancer or existence of infectious diseases.

**Table 1.**
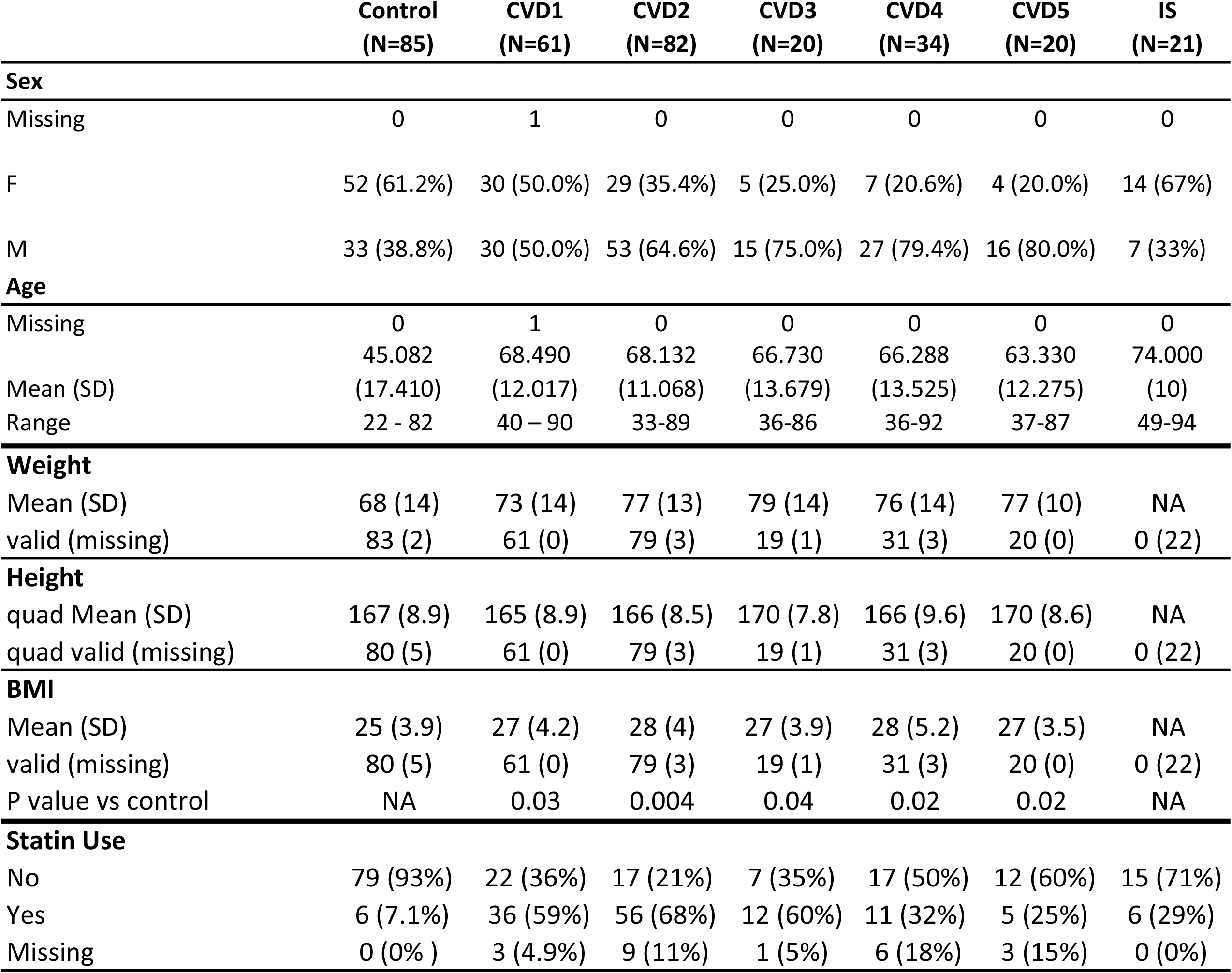
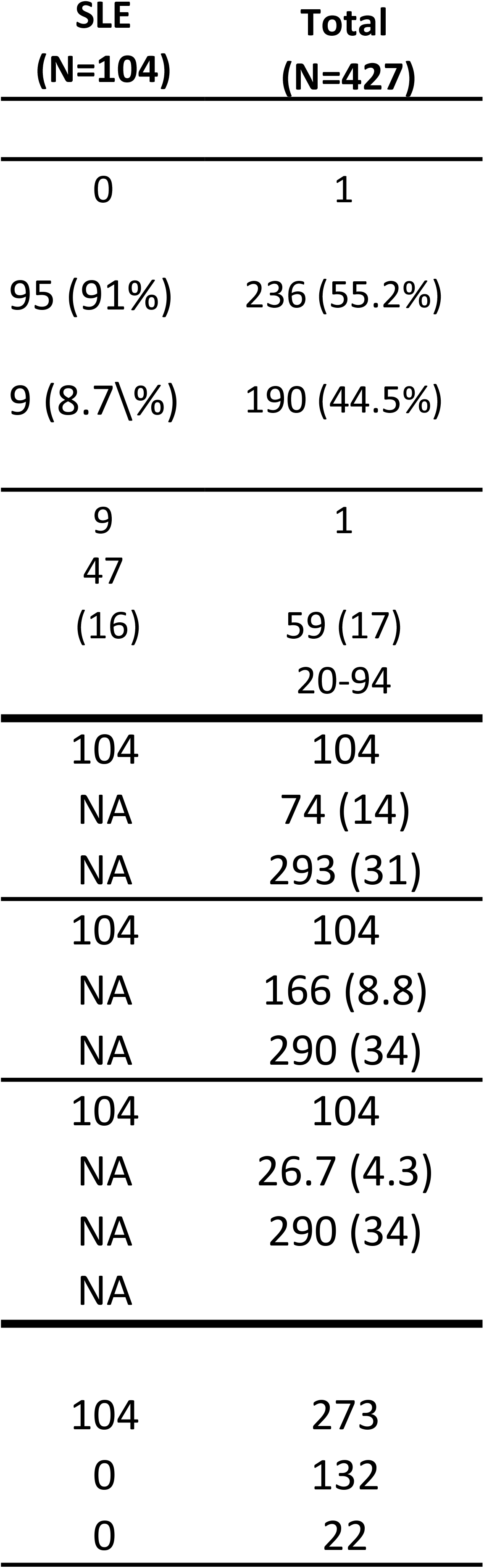
Baseline characteristics of the analyzed lipid cohort.

Plasma was obtained from all donors after explaining the purpose and obtaining written informed-consent from them or their legal representatives. The entire process was approved by the Ethical Review Board of the Faculty of Medicine of the New University of Lisbon and the Ethics Committee for Health of the Centro Hospitalar de Lisboa Ocidental, that includes the Hospital Santa Cruz, the Hospital Egas Moniz and Hospital São Francisco Xavier, and the Ethics Committee for Health of the Hospital Fernando Fonseca. All experiments were performed in accordance with the guidelines and regulations. Blood samples were drawn into tubes containing an anti-coagulant (heparin or EDTA) immediately after admission into the hospital and signing of the informed consent. The samples were kept at 4°C and processed within 24 h from collection. Plasma was obtained by centrifugation of the blood at 500 g for 10 min at 4°C, frozen at −80°C and stored at this temperature until they were used for the lipidomic analysis.

### Definition of age matched controls

The total control cohort consisted of people with ages from 22 years to 82 years. Of these only those with ages ≥36 years (n = 52) were used for comparison with the CVD and IS cohorts. The whole control cohort (n = 85) was used for comparison with SLE patients.

### Lipid extraction for mass spectrometry lipidomics

Mass spectrometry-based lipid analysis was performed at Lipotype GmbH (Dresden, Germany) as described [37]. For lipid extraction an equivalent of 1 μL of undiluted plasma was used. Internal standards were pre-mixed with the organic solvents mixture and included: cholesterol D6, cholesteryl ester 20:0, ceramide 18:1;2/17:0, diacylglycerol 17:0/17:0, phosphatidylcholine 17:0/17:0, phosphatidylethanolamine 17:0/17:0, lysophosphatidylcholine 12:0, lysophosphatidylethanolamine 17:1, triacylglycerol 17:0/17:0/17:0, and sphingomyelin 18:1;2/12:0. All liquid handling steps were performed using Hamilton Robotics STARlet robotic platform with the Anti Droplet Control feature for organic solvents pipetting.

### MS data acquisition

Samples were analyzed by direct infusion in a QExactive mass spectrometer *(Thermo* Scientific) equipped with a TriVersa NanoMate ion source (Advion Biosciences). Samples were analyzed in both positive and negative ion modes with a resolution of R_m/z=200_ = 280000 for MS and R_m/z=200_ = 17500 for MSMS experiments, in a single acquisition. MSMS was triggered by an inclusion list encompassing corresponding MS mass ranges scanned in 1 Da increments. Both MS and MSMS data were combined to monitor CE, DAG and TAG ions as ammonium adducts; PC, PC O-, as acetate adducts; and PE, PE O- and PI as deprotonated anions. MS only was used to monitor LPE as deprotonated anion; Cer, SM and LPC as acetate adducts and cholesterol as an ammonium adduct.

### Lipid nomenclature

The following annotations were used: Lipid class-<sum of carbon atoms>:<sum of double bonds>;<sum of hydroxyl groups>, *i*.*e*. SM-34:1;2 means an SM lipid with 34 carbon atoms, 1 double bond and 2 hydroxyl groups in the ceramide backbone. Lipid molecular subspecies annotation [38] contains additional information on the exact identity of their fatty acids. For example PC 18:1;0_16:0;0 denotes a phosphatidylcholine with one acyl chain having 18 carbon atoms, 1 double bond, 0 hydroxylation, and a second acyl chain with 16 carbon atoms, o double bonds, 0 hydroxylation. The exact position of the fatty acids in relation to the glycerol backbone (*sn*-1 or *sn*-2) cannot be discriminated. CE 18:1;0 denotes a cholesteryl ester with an 18:1;0 fatty acid. Lipid identifiers of the SwissLipids database [38] are provided in the supplemental dataset.

### Post-processing

Data were analyzed with in-house developed lipid identification software based on LipidXplorer [39, 40]. Data post-processing and normalization were performed using an in-house developed data management system. Only lipid identifications with a signal-to-noise ratio >5, and a signal intensity 5-fold higher than in corresponding blank samples were considered for further data analysis. Using 8 reference samples per 96-well plate batch, lipid amounts were corrected for batch variations. Amounts were also corrected for analytical drift, if the p-value of the slope was below 0.05 with an R^2^ greater than 0.6 and the relative drift was above 5%. Median coefficient of (sub-) species variation as accessed by reference samples was 3.1%. The full data set contained quantitative information from 623 lipids. Lipids with a concentration less than 0.5 μM were considered “not analyzable” (NA) and, for the purposes of the present work were considered to be at half of the minimum detectable value and zero variance lipids were filtered out providing a data set of 596 lipids which served as the input to subsequent multivariate analyses.

### Significant regulated lipids

were defined by the R package “limma version 3.42.0”. The data obtained by MS were added one and log2 transformed. Next the quantitative values were normalized across samples using robust quantile normalization [41]. Both the raw and the quantile normalized values were analyzed by the R package “limma version 3.42.0” to determine significant regulated lipids between control and patient cohorts. Linear models included terms to correct for gender and statin treatment and only age matched controls were included for statistical regulation analysis involving IS and CVD cases. All controls were included for comparison with the SLE group. SLE group was sub divided into two groups according to the Systemic Lupus International Collaborating Clinics (SLICC) group criteria (SLICC >=4 versus SLICC < 4) [42]. Statistical differential regulated lipids were determined using the R package “limma version 3.42.0” and p values were adjusted for multiple testing by the method of Benjamini and Hochberg [43]. Principal component analysis was performed using R base functions on centered quantile normalized quantification values. Overlap between significant regulated lipids was performed with the R package “VennDiagram”.

### Supervised classification

Linear discriminant analysis (LDA) was performed using the R package “caret version 6.0.84” [44] as interface to “MASS version 7.3.51.4”. LDA was performed using either all 596 zero variance filtered lipids or 206 lipids selected with more than 50% non NAs per clinical group. Additionally, the parameters for training and testing of LDA were set to remove lipids with total intensity across all samples below 30. The value 30 was obtained by stepwise optimization to establish best possible separations. The correlation cut off was set to 1 which means that correlated variables were not eliminated (caret parameter pair-wise absolute correlation cutoff). Setting the correlation cut off slightly lower, e.g. 0.9-1 had no effect of on the separation in the LDA plots. The results from the two approaches resulted in similar classification performance and for simplicity only the results from the 596 zero variance filtered lipids is presented.

Partial least square (PLS) analysis was performed using the R package “caret version 6.0.84” [44] as interface to “pls version 2.7.1“. The full data set 427 individuals were split into disease groups with age matched controls. By random, 75% of the data from smallest groups were used for training the PLS model and balanced age matched samples from the majority group sampled to create balanced training sets. The remaining data were left out and used to obtain the final accuracy measures (validation set). The caret preprocessing parameters specified were: “zv – exclude zero variance predictors, “center – subtracts the mean from predictor values”, and “scale – divide predictor values with the standard deviation”. The model was optimized by 10 fold cross validation repeated 10 times using accuracy as the metric for optimization. ROC curves and area under the curve were estimated using the R package “pROC” [45] for model performance evaluation on the left out validation set.

## Results

### Baseline characteristics

Table 1 summarizes the baseline characteristics. The CVD cohort analyzed in this study consisted of 217 participants with mean age of 67 years (range 33-92 years). A total of 21 IS patients were included with a similar age range as the CVD patients. Statistical comparisons were in all cases performed with aged-matched controls. The entire control cohort (n = 85) consisted of people with ages between 22 and 82 years. Of this group a sub-group (n = 52) with ages ≥36 years served as the control for comparisons with the CVD + IS, while the entire control cohort (n = 85) was used for comparison with the SLE patients.

### Lipidomic analysis

Based on a single shot MS based lipid analysis of ten microliters of plasma, 623 lipids spanning 15 lipid classes were identified and quantified. The analysis presented in the current study used only 596 of these lipids and demonstrated, as discussed below, that accurate classification of SLE, IS, CVD and CVD sub-groups from controls was achievable in spite of the fact that there were several shared dysregulated lipids among the different cohorts.

### Unsupervised analysis

Principal component analysis based on all 596 selected lipids provided a reasonable separation between aged-matched controls versus CVD1, CVD2, CVD3, CVD4, CVD5 and IS (Figure 1A-F). CVD4 and CVD5 are almost fully separable from controls based on the first two principal components. CVD3 versus control resulted in the poorest separation by the first two principal components. Depending on the condition either PC1 or PC2 provided the most separation. This strongly suggests that the largest or second largest variance component in lipid abundance correlate with these disease conditions. Notably, this PCA analysis was performed using quantitative values from all lipids without any biased pre-selection of lipids known to associate to CVD and IS.

**Figure 1.**
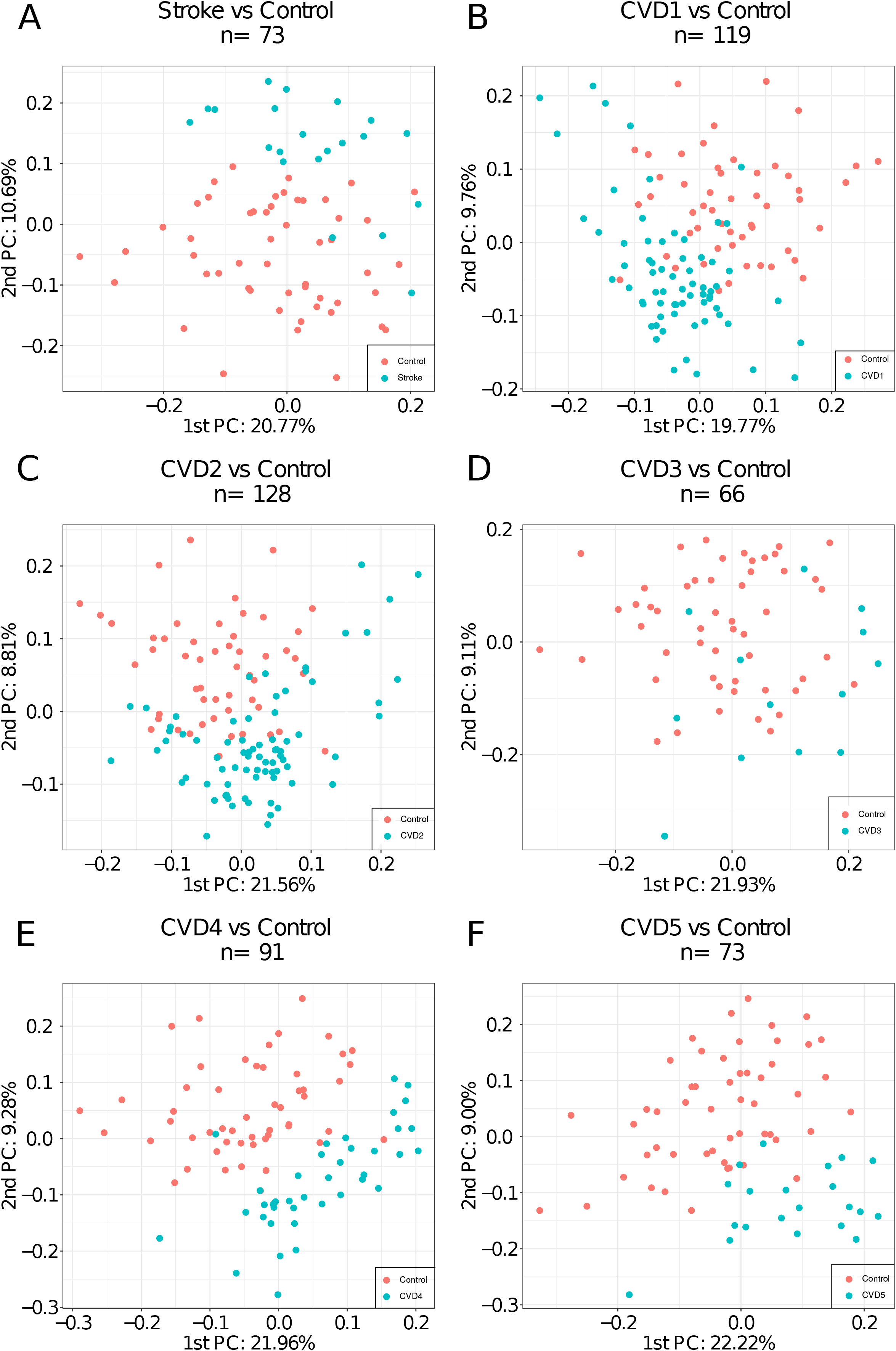
Principal component analysis using all 596 selected lipids as input. Plot of the first and second principal component for A) IS, B) CVD1, C) CVD2, D) CVD3, E) CVD4, and F) CVD5.

### Association of lipids with clinical diagnosis

Comparison of SLE sub-groups SLE_SLICC criteria ≥ 4 vs < 4 resulted in no significant lipids after correction of multiple testing (Table S2). Pairwise comparison between age matched controls and the patient groups adjusted for gender and statin use revealed the following number of significant dysregulated lipids after correction of multiple testing (First value: P adjusted <0.05/second value: P adjusted <0.05 and at least two-fold regulated, Table S2): 147/13 CVD1, 158/17 CVD2, 78/16 CVD3, 130/21 CVD4, 105/19 CVD5, 98/11 IS and 166/108 SLE. Among controls the number of significant regulated lipids after correction of multiple testing (P adjusted <0.05/P adjusted <0.05 and at least two-fold regulated) were 26/8 when comparing controls with and without statin treatment (Table S2). Only about 10-20% of the significant (dys)regulated lipids displayed an effect size bigger or equal to two-fold, prompting the question if the lipids with small effect size are diagnostically relevant.

Therefore, Venn diagrams were used to compare the overlap of significant dysregulated lipids (Figure 2A) and significant dysregulated lipids with an effect size more than two-fold (Figure 2B). We observed that a considerable number of significant dysregulated lipids with effect size less than two-fold were shared across pathological groups (Figure 2AB). Heatmaps were constructed to address the direction of the shared dysregulated lipids (Figure 2CD). These heatmaps only depict lipids that are regulated in more than one pairwise comparison. The direction of dysregulation of lipids displays large similarities for the CVD, IS and SLE groups. In contrast, the lipids found regulated between controls and controls treated with statin displayed a reverse pattern in the direction of regulation compared to the pathological groups (Figure 2CD). Next, the frequency of lipid classes of the significant regulated lipids for pathological groups versus statin regulation were compared (Figure 3ABCD). Predominantly the lipid classes PC, CE, SM including cholesterol were up-regulated in the pathological conditions (Figure 3A) which to some extent match the statin down-regulation of CE, PC and cholesterol (Figure 3D). Similarly, the down regulation of lipid classes TAG, PC, PE and DAG in pathological conditions match, to some degree, the observed up-regulation of TAG, DAG and PE for statin treated patients. In general, pairwise comparison for disease groups or disease sub-groups versus control resulted in statistical differences in lipid expression (table S2). This was also the case when different disease groups were compared pairwise against each other. However, sub-comparisons of CVD (e.g. CVD1 versus CVD5) or SLE (e.g. SLICC ≥ 4 versus SLICC < 4) resulted in fewer significant regulated lipids after correcting for multiple testing. Especially, SLICC ≥ 4 versus SLICC < 4 comparison resulted in no significant regulated lipids after correction for multiple testing.

**Figure 2.**
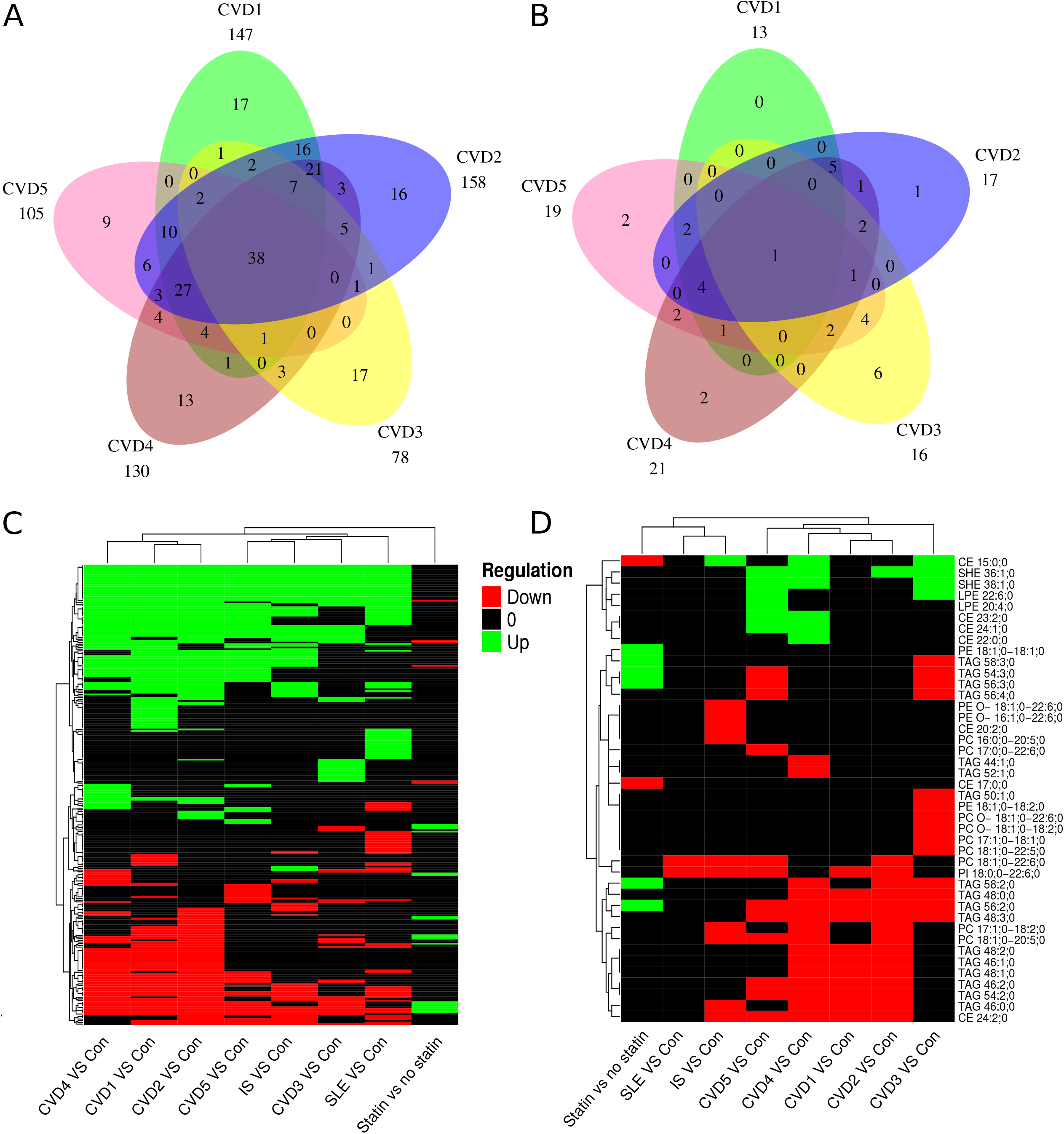
Significant regulated lipids across comparisons. Venn comparison of significant regulated lipids after correction of multiple testing (A) and with two fold regulation (B) for CVD groups. C) Heatmap depicting the direction of regulation for significant regulated lipids across comparisons. D) Heatmap depicting the direction of regulation for significant regulated lipids with more than two fold regulation across comparisons.

**Figure 3.**
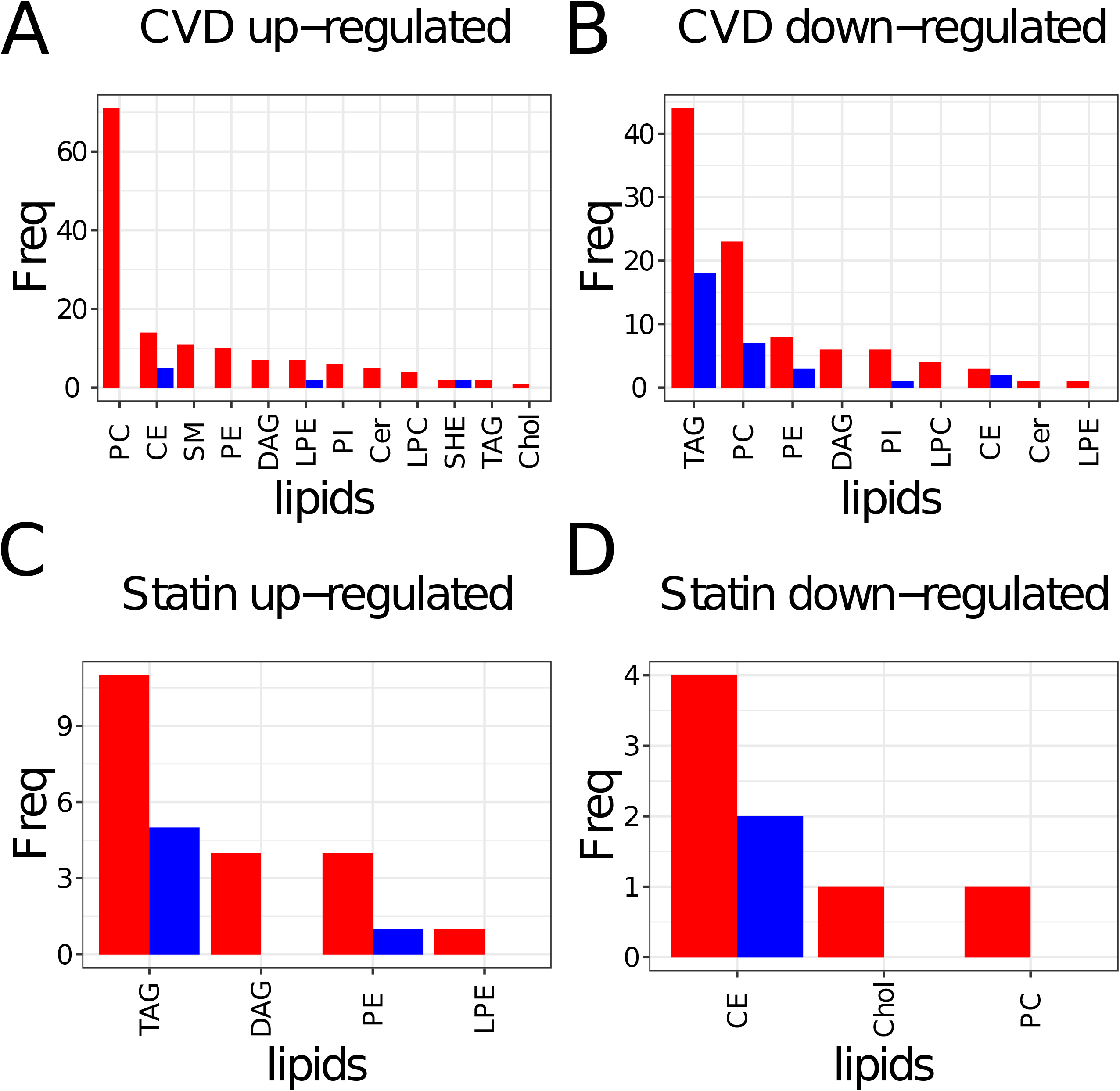
Frequency of lipid classes among significant regulated lipids. Significant up (A) and down (B) regulated lipid classes for CVD versus control. Significant up (C) and down (D) regulated lipid classes for statin treated versus control. Red bars indicate significant regulated lipid class frequencies after correction of multiple testing. Blue bars indicate significant regulated lipid class frequencies after correction of multiple testing and filtering for at least two fold regulation.

### Supervised analysis

Preliminary linear discriminant analysis (LDA) based on the 596 selected lipids and all data displayed strong potential for building classifiers for separating the pathological groups and control (Figure 4). Even the individual CVD sub-groups were fairly effectively separated in the LDA plots (Figure 4AB). Analyzing the CVD groups and controls separately further supported the potential of lipids to stratify CVD severity groups (Figure 4B). LDA separates SLE from controls with a P value < 2.2e-16 (Figure 4C). Figure 4D depicts the LDA plot for controls versus IS. The first LDA component significantly separates IS patients from controls (P value < 2.4e-13). The PCA (Figure 1) and LDA (Figure 4) plots encouraged us to build and test partial least square models (PLS). As a proof of concept three PLS classifiers with high classification performance when tested on an independent data set were established (Figure 5).

**Figure 4.**
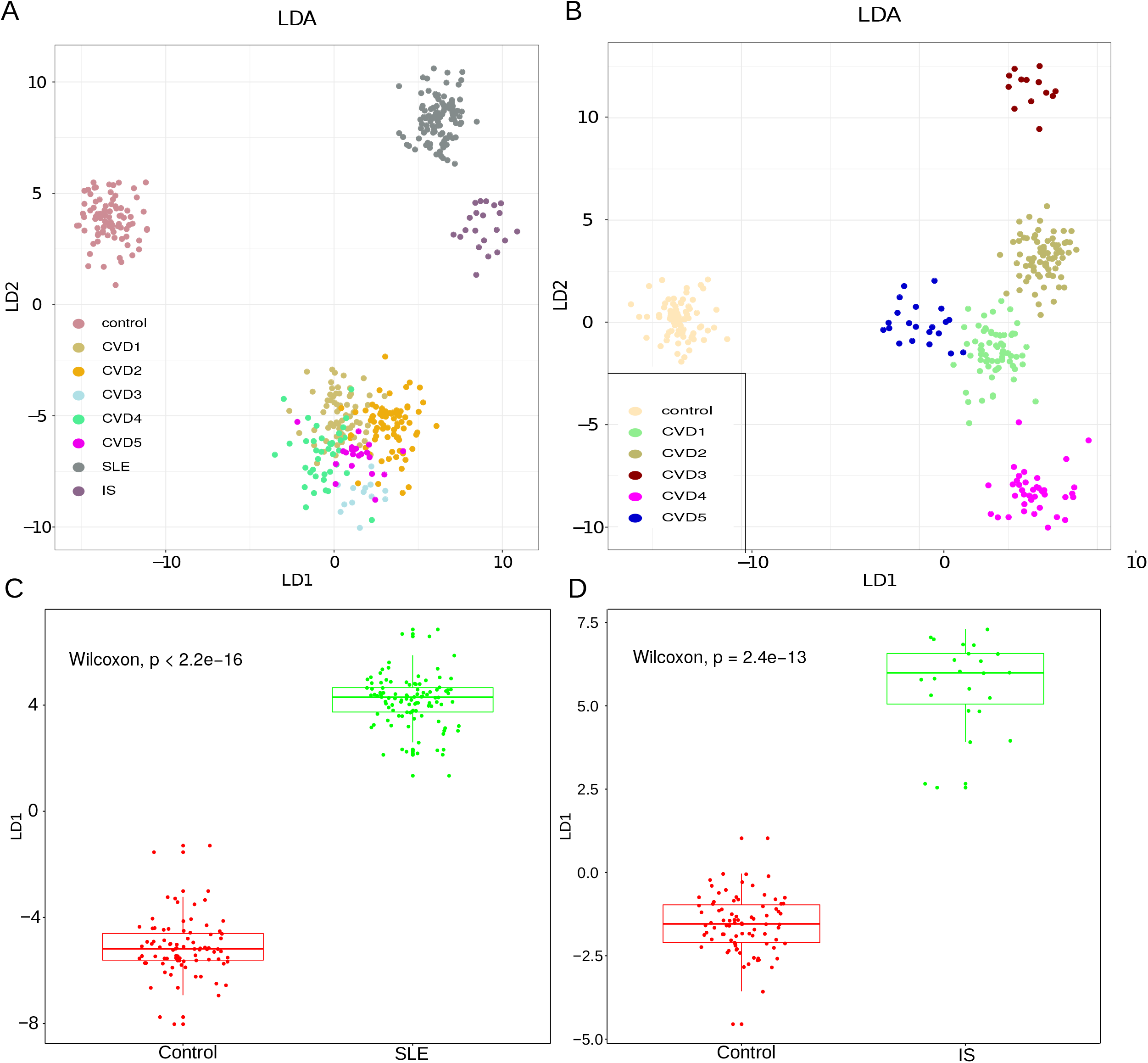
Separation of the disease groups by LDA based on selected lipids. A) LDA plots demonstrating separation of CVD, SLE, IS and control. B) LDA base separation of CVD groups and control samples. C) First LDA component versus SLE and control. D) First LDA component versus IS and control.

**Figure 5.**
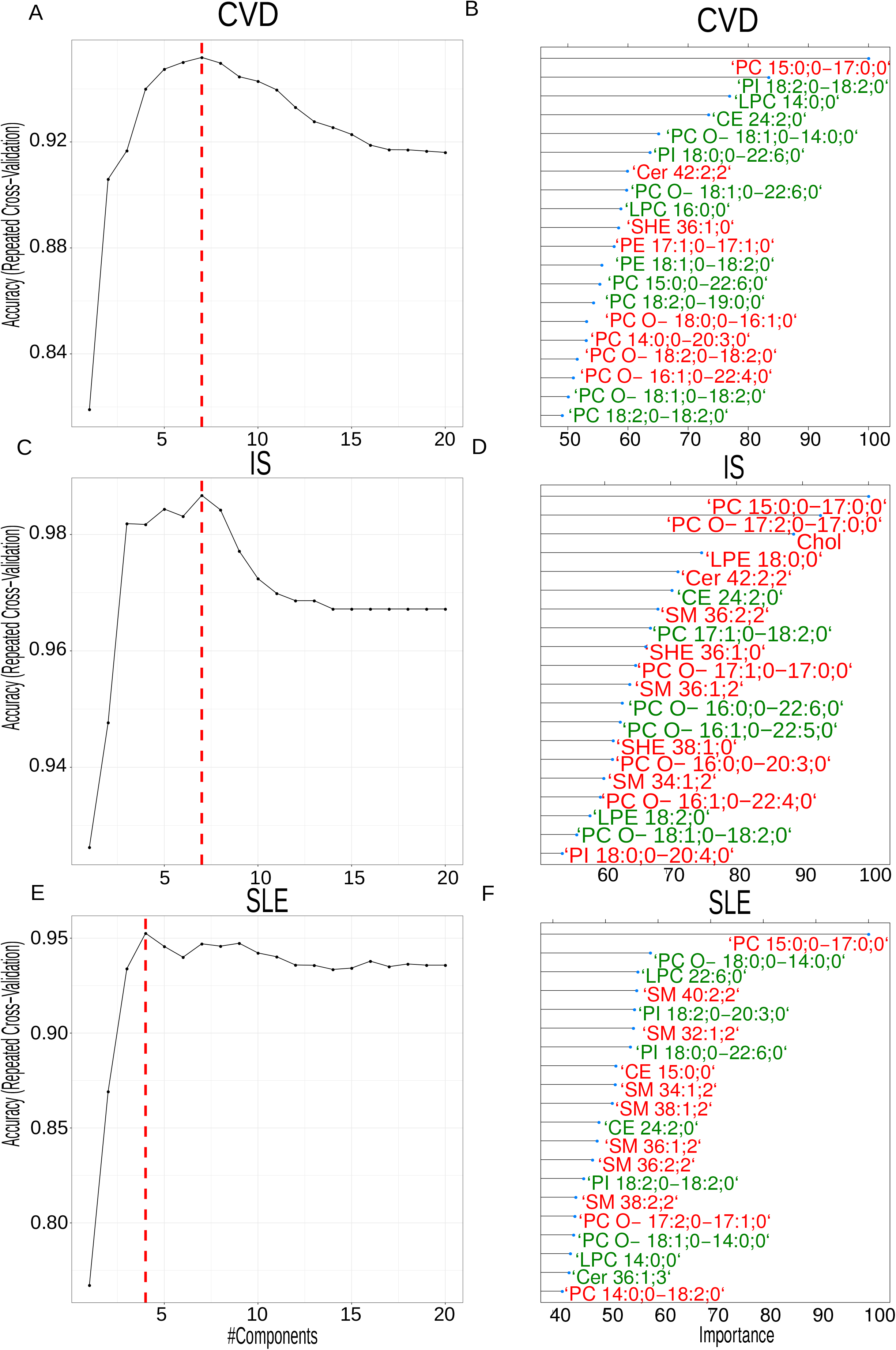
Training of PLS models. Left panel: Optimal number of PLS components for model based on A) CVD versus control, C) IS versus control, and E) SLE versus control. Right panel (BDF): Indicate the 20 most important lipids in the PLS models. Red labels indicate overall higher abundance in disease whereas green labels indicate overall higher abundance in controls.

The optimal number of PLS component for separation of CVD and controls was seven (Figure 5A). The right panel displays the 20 most important lipids contributing to the PLS components. The width of each bar is indicative of the lipid’s importance in the model. The optimal PLS component for IS versus control were seven (Figure 5C) and the 20 highest ranked lipids in terms of importance are depicted in the right panel. For SLE versus controls four PLS components were optimal (Figure 5E). The 20 most important lipids for SLE displayed some similarity to the most relevant lipids for CVD versus control and IS versus control (Figure 5BDF). For example, “PC 15:0;0_17:0;0” were low abundant in controls compare to SLE, IS and CVD (Table S2). The three PLS models for separating IS, CVD and SLE from age matched controls was validated by using a validation set left out from the training and optimization of the PLS models (Figure 6). The ROC curve for classification performance of CVD versus controls is depicted in Figure 6A. The area under the curve was 0.99. The confusion matrix is inserted in the lower right side of figure 6A and indicates that 72 out of 75 of the left-out data set were correctly classified. The model for CVD1 versus controls displayed a slightly worse classification performance with five out of 52 misclassified (Accuracy ≈90%, area under the curve was 0.97). Although, linear discriminant analysis based on all data displayed partial to complete separation for CVD1 versus CVD2 (Figure 4AB) the PLS model was not much better than random when tested on the independent data set (area under the roc curve=0.53).

**Figure 6.**
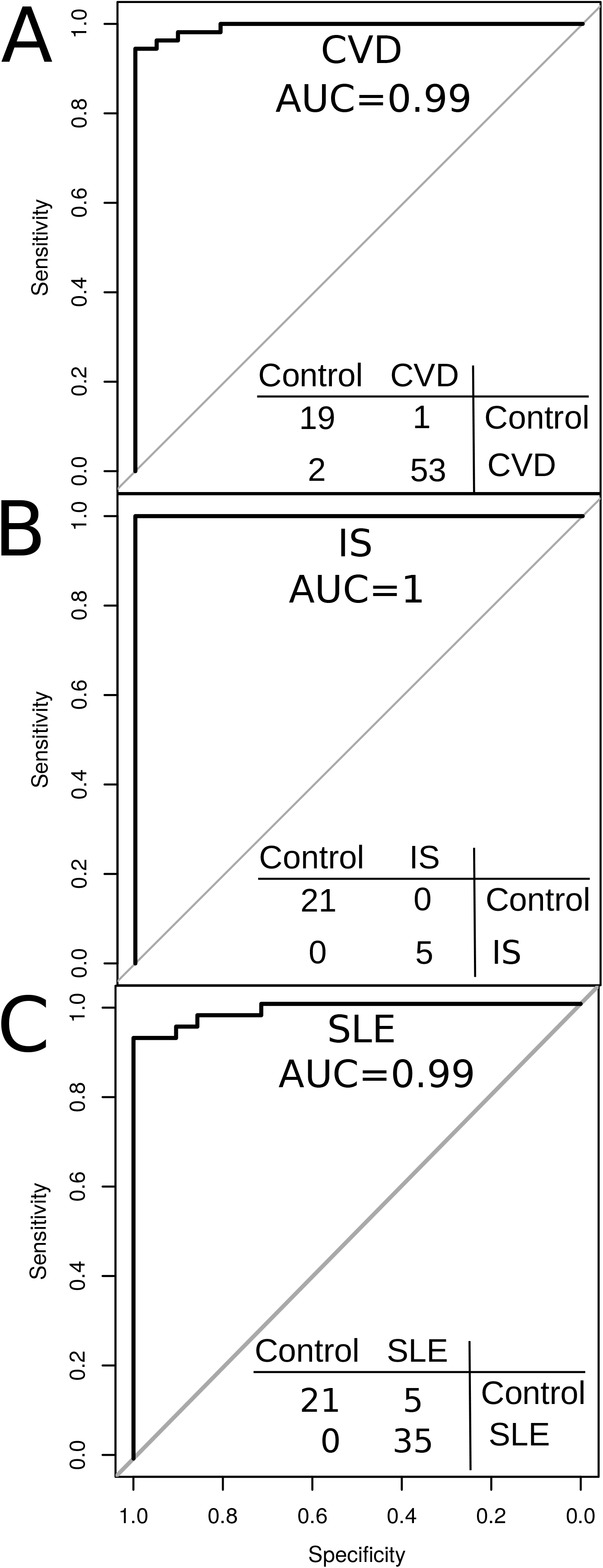
Classification performance illustrated with ROC curves and confusion matrix for A) CVD versus control, B) IS versus control, and C) SLE versus control.

The ROC curve for the PLS model for classification of IS versus controls when tested on the independent validation data set is depicted in figure 6B. The IS model misclassified zero patients in the independent validation out of total of 26 samples (Accuracy=100%). The ROC curve for the PLS model for classification of SLE versus controls displayed an area under the curve of 1 and no misclassified subjects. The PLS model for SLE versus control demonstrated similar classification performance as CVD versus controls with a total of five misclassified cases out of 61 (Figure 6C).

In conclusion, we present the accuracy of separation, via statistical analysis of the lipidomic data, of the cohorts studied – namely, control, SLE, IS, and CVD1 through CVD5. The accuracy of separating SLE and CVD cases on an independent test data set was above 91%. The accuracy of separating IS versus SLE and IS versus CVD was 78%. Figure 7 summarizes pairwise classification accuracies for all CVD subgroups versus controls when evaluated on an independent teat data set. All CVD cases were separated from the controls with accuracies above 80%. Pairwise classifications of CVD1 versus CVD2 and CVD4 versus CVD5 resulted in an accuracy which was only slightly better than random. These pairwise classification accuracies are concurrent with the number of significantly identified lipids in the pairwise comparisons obtained from the linear regression models (Table S2). Overall, these findings suggest that plasma lipidomics profiles have the potential to accurately distinguish chronic inflammatory diseases from controls and that the lipidomic profiles are characteristics of the pathophysiological states.

**Figure 7.**
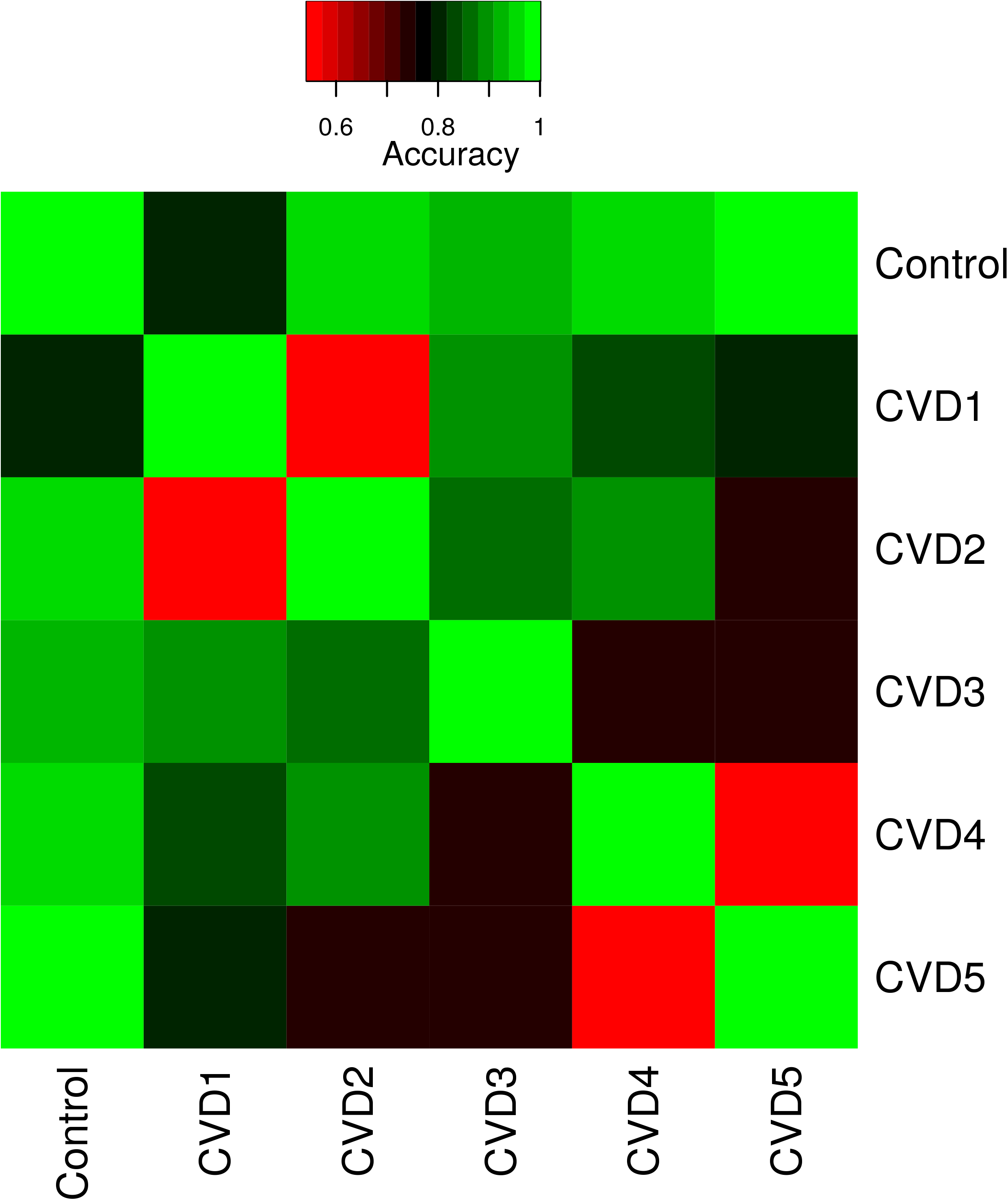
Summary of classification performance for all pairwise comparison of control and CVD1 to CVD5. The heatmap depicts the classification accuracy obtained on an independent test set for each of the pairwise comparisons.

## Discussion

Blood plasma is the medium through which the physiological steady state of lipid distribution in the body is maintained. These lipids may be of dietary or auto-synthetic origin but are also the result of degradative chemical processes (oxidation, lysis, modification in the blood plasma, etc.) and cell death due to acute or chronic processes such as inflammation. The exact steady state concentration of each lipid in the plasma is therefore a combination of dietary, physiological-chemical, genetic, and patho-physiological states. Many lipidic products are known to positively or negatively influence inflammatory processes. Lipidomics may therefore hold a promise as a valuable tool for identifying and distinguishing between different patho-physiological states. There are perhaps well over a thousand different lipids and their derivative products in the blood plasma. Therefore, the more lipids are identified and quantified, the better will be our capacity to diagnose the physiological state. This view seems to be reinforced by recent developments [46].

We have, therefore, used lipidomic data on three separate cohorts – a cohort consisting of people who suffer from SLE, a cohort of people who suffer from CVD (including IS), and an age-matched Control cohort characterized by the fact that they never sought medical help for any of the disease states of the other two cohorts. The CVD cohort consisted of five patient groups, CVD-1 through CVD-5, as described earlier. The patients of IS, SLE and CVD have a common characteristic – chronic inflammation – but distinct primary etiologies. The comparison of these two patho-physiological states therefore provides an efficient way to evaluate the ability of lipidomics to distinguish between them and possibly serve eventually as a diagnostic tool. A total of 596 lipids divided into 15 lipid classes were analyzed using shotgun lipidomics and adequate statistical methods were used to analyze the results. We note that our aim has not been to identify the lipids characteristic of one pathological state or another, although some conclusions in this regard may also be drawn, but rather to use the data to create distinct diagnostic groups that correspond with the distinct pathological states.

The improvement in lipid identification and quantification from plasma samples presented in this study led to improved classification performance of SLE, CVD and CVD sub-groups, and IS from controls. This reinforces the potential of plasma lipidomic profiles as biomarkers for cardiovascular risk stratification proposed in previous studies [16]. We observed clear commonalities between dysregulated lipids in CVD and in IS and to a lesser extent in SLE (Figure 2CD). This observation supports the idea of a common etiology for CVD and IS which involve atherosclerotic lesions in blood vessels in both cases. It further strengthens the link between SLE and CVD as CVD is overrepresented in patients with SLE and SLE patients demonstrate accelerated atherosclerosis [47-50]. The reverse regulation observed for lipids in statin treated versus CVD and IS suggests that statin to some extent stabilize the lipid profiles towards lipid profiles of disease-free controls. Nevertheless, the most frequent regulated lipid class in CVD and stroke were only moderately affected by statin (Figure 4A versus 4D). The large number of dysregulated PCs does not appear affected by statin treatment and may constitute attractive pharmaceutical targets for future treatment modalities. CVD1 constitutes patients who complain about chest pain but no pathological diagnostic indicators for CVD were identified. The trained classifiers on lipid abundance were, however, able to distinguish between control and CVD1. However, it must be noted that the classification between CVD1 and CVD2 were basically close to random. This suggests that lipid profiling is the first clinical indicator that can diagnose CVD1 individuals. This finding may have both therapeutic and diagnostic impact and must be further explored in future studies.

In conclusion, we present the accuracy of separation on independent lipidomics test data, of the cohorts studied – namely, control, SLE, IS, and CVD1 through CVD5. The accuracy of separating SLE and CVD cases was above 0.91. The accuracy of separating IS versus SLE and IS versus CVD was 0.78. Figure 7 summarizes pairwise classification accuracies for all CVD subgroups versus controls when evaluated on an independent test data set. All CVD cases were separated from the controls with accuracies above 0.80. The high accuracies obtained in our study may result from the higher number of lipids profiled. However, the cohorts used were not ideal in terms of sex distribution across CVD conditions and controls. Furthermore, the age matched controls had a slightly lower average age than the CVD and stroke groups. No statistical differences between the controls groups and CVD groups for BMI was identified (Table 1). The samples for each condition were also not obtained from multiple centers which may introduce bias.

Pairwise classifications of CVD1 versus CVD2 and CVD4 versus CVD5 resulted in an accuracy which was only slightly better than random. These pairwise classification accuracies are concurrent with the number of significantly identified lipids in the pairwise comparisons obtained from the linear regression models (Table S2). Overall, these findings suggest that plasma lipidomics profiles have the potential to accurately distinguish chronic inflammatory diseases from controls and that the lipidomic profiles are characteristics of the pathophysiological states. The necessary criterion is that the lipidomic data contain as many lipids as possible. We suggest that, given the ease of shotgun lipidomic quantification of a very large number of lipids in blood plasma and the high accuracy of the separation and identification of chronic inflammatory pathologies upon analysis of the lipidomic data, the methods described in this work could be a valuable tool in early diagnostic methodology.

### Study limitations

The study is limited by the number of samples per group especially when subdividing into cardiovascular subgroups. Gender balance is unbalanced across stroke and cardiovascular disease (see Table 1). Women were represented higher in the ischemic stroke groups whereas men were more represented in subsets of the cardiovascular groups. Age distribution for controls was moderately lower than cardiovascular and IS group. The reverse regulation of lipids when comparing statin treated controls with CVD and IS is an explorative result and the observed regulation might originate due to confounding factors. This observation should preferably be validated in a randomized control study.

We understand the present study as an exploratory preliminary study and clearly more inflammatory diseases need to be compared. Recommendations with regard to diagnostics will require much larger cohorts with better matching of age, sex, statin use, BMI, etc. But this will be a process that will require many more years and preferably collaborative work between many laboratories.

## Data Availability

All data referred to in the manuscript will be available

## ACKNOWLEDGEMENTS

We thank the nursing staff at the Hospital Santa Cruz, in particular Edite Tomás Mateus and Manuel Belo Costa for their enthusiastic involvement in this project and help in blood collection.

---

**Table S1**. Significant regulated lipids and trends found in similar studies and discussed in the introduction.

**Table S2**. Quantitative lipid data and statistical outcome based on limma analysis applying linear models correcting for gender and statin treatment.

## References

1. Hansson, G.K., A.K. Robertson, and C. Soderberg-Naucler, Inflammation and atherosclerosis. Annu Rev Pathol, 2006. 1: p. 297–329.

2. Weber, C. and H. Noels, Atherosclerosis: current pathogenesis and therapeutic options. Nat Med, 2011. 17(11): p. 1410–22.

3. Stegemann, C., et al., Comparative lipidomics profiling of human atherosclerotic plaques. Circ Cardiovasc Genet, 2011. 4(3): p. 232–42.

4. Ding, M. and K.M. Rexrode, A Review of Lipidomics of Cardiovascular Disease Highlights the Importance of Isolating Lipoproteins. Metabolites, 2020. 10(4).

5. Nelson, J.C., et al., Plasma sphingomyelin and subclinical atherosclerosis: findings from the multi-ethnic study of atherosclerosis. Am J Epidemiol, 2006. 163(10): p. 903–12.

6. Fernandez, C., et al., Plasma lipid composition and risk of developing cardiovascular disease. PLoS One, 2013. 8(8): p. e71846.

7. Diaz, S.O., et al., Exploratory analysis of large-scale lipidome in large cohorts: are we any closer of finding lipid-based markers suitable for CVD risk stratification and management? Anal Chim Acta, 2021. 1142: p. 189–200.

8. Sigruener, A., et al., Glycerophospholipid and sphingolipid species and mortality: the Ludwigshafen Risk and Cardiovascular Health (LURIC) study. PLoS One, 2014. 9(1): p. e85724.

9. Stegemann, C., et al., Lipidomics profiling and risk of cardiovascular disease in the prospective population-based Bruneck study. Circulation, 2014. 129(18): p. 1821–31.

10. Tarasov, K., et al., Molecular lipids identify cardiovascular risk and are efficiently lowered by simvastatin and PCSK9 deficiency. J Clin Endocrinol Metab, 2014. 99(1): p. E45–52.

11. Cheng, J.M., et al., Plasma concentrations of molecular lipid species in relation to coronary plaque characteristics and cardiovascular outcome: Results of the ATHEROREMO-IVUS study. Atherosclerosis, 2015. 243(2): p. 560–6.

12. Laaksonen, R., et al., Plasma ceramides predict cardiovascular death in patients with stable coronary artery disease and acute coronary syndromes beyond LDL-cholesterol. Eur Heart J, 2016. 37(25): p. 1967–76.

13. Alshehry, Z.H., et al., Plasma Lipidomic Profiles Improve on Traditional Risk Factors for the Prediction of Cardiovascular Events in Type 2 Diabetes Mellitus. Circulation, 2016. 134(21): p. 1637–1650.

14. Havulinna, A.S., et al., Circulating Ceramides Predict Cardiovascular Outcomes in the Population-Based FINRISK 2002 Cohort. Arterioscler Thromb Vasc Biol, 2016. 36(12): p. 2424–2430.

15. Wang, M., et al., Plasma 7-ketocholesterol levels and the risk of incident cardiovascular events. Heart, 2017. 103(22): p. 1788–1794.

16. Mundra, P.A., et al., Large-scale plasma lipidomic profiling identifies lipids that predict cardiovascular events in secondary prevention. JCI Insight, 2018. 3(17).

17. Razquin, C., et al., Plasma lipidome patterns associated with cardiovascular risk in the PREDIMED trial: A case-cohort study. Int J Cardiol, 2018. 253: p. 126–132.

18. Harshfield, E.L., et al., An Unbiased Lipid Phenotyping Approach To Study the Genetic Determinants of Lipids and Their Association with Coronary Heart Disease Risk Factors. J Proteome Res, 2019. 18(6): p. 2397–2410.

19. Anroedh, S., et al., Plasma concentrations of molecular lipid species predict long-term clinical outcome in coronary artery disease patients. J Lipid Res, 2018. 59(9): p. 1729–1737.

20. Wang, D.D., et al., Plasma Ceramides, Mediterranean Diet, and Incident Cardiovascular Disease in the PREDIMED Trial (Prevencion con Dieta Mediterranea). Circulation, 2017. 135(21): p. 2028–2040.

21. Meikle, P.J., et al., Plasma lipidomic analysis of stable and unstable coronary artery disease. Arterioscler Thromb Vasc Biol, 2011. 31(11): p. 2723–32.

22. Ellims, A.H., et al., Plasma lipidomic analysis predicts non-calcified coronary artery plaque in asymptomatic patients at intermediate risk of coronary artery disease. Eur Heart J Cardiovasc Imaging, 2014. 15(8): p. 908–16.

23. Bellis, C., et al., Human plasma lipidome is pleiotropically associated with cardiovascular risk factors and death. Circ Cardiovasc Genet, 2014. 7(6): p. 854–863.

24. Ramo, J.T., et al., Coronary Artery Disease Risk and Lipidomic Profiles Are Similar in Hyperlipidemias With Family History and Population-Ascertained Hyperlipidemias. J Am Heart Assoc, 2019. 8(13): p. e012415.

25. Yin, X., et al., Lipidomic profiling identifies signatures of metabolic risk. EBioMedicine, 2020. 51: p. 102520.

26. Gerl, M.J., et al., Cholesterol is Inefficiently Converted to Cholesteryl Esters in the Blood of Cardiovascular Disease Patients. Sci Rep, 2018. 8(1): p. 14764.

27. Zhang, T. and C. Mohan, Caution in studying and interpreting the lupus metabolome. Arthritis Res Ther, 2020. 22(1): p. 172.

28. Ferreira, H.B., et al., Lipidomics in autoimmune diseases with main focus on systemic lupus erythematosus. J Pharm Biomed Anal, 2019. 174: p. 386–395.

29. Shah, D., et al., Oxidative stress and its biomarkers in systemic lupus erythematosus. J Biomed Sci, 2014. 21: p. 23.

30. Reichlin, M., et al., Autoantibodies to lipoprotein lipase and dyslipidemia in systemic lupus erythematosus. Arthritis Rheum, 2002. 46(11): p. 2957–63.

31. Borba, E.F., et al., Chylomicron metabolism is markedly altered in systemic lupus erythematosus. Arthritis Rheum, 2000. 43(5): p. 1033–40.

32. Hu, C., et al., Oxidative stress leads to reduction of plasmalogen serving as a novel biomarker for systemic lupus erythematosus. Free Radic Biol Med, 2016. 101: p. 475–481.

33. Lu, L., et al., Shotgun Lipidomics Revealed Altered Profiles of Serum Lipids in Systemic Lupus Erythematosus Closely Associated with Disease Activity. Biomolecules, 2018. 8(4).

34. Fihn, S.D., et al., 2012 ACCF/AHA/ACP/AATS/PCNA/SCAI/STS Guideline for the Diagnosis and Management of Patients With Stable Ischemic Heart Disease: Executive Summary: A Report of the American College of Cardiology Foundation/American Heart Association Task Force on Practice Guidelines, and the American College of Physicians, American Association for Thoracic Surgery, Preventive Cardiovascular Nurses Association, Society for Cardiovascular Angiography and Interventions, and Society of Thoracic Surgeons. J Am Coll Cardiol, 2012. 60(24): p. 2564–603.

35. Thygesen, K., et al., Fourth Universal Definition of Myocardial Infarction (2018). Circulation, 2018. 138(20): p. e618–e651.

36. Amsterdam, E.A., et al., 2014 AHA/ACC guideline for the management of patients with non-ST-elevation acute coronary syndromes: a report of the American College of Cardiology/American Heart Association Task Force on Practice Guidelines. Circulation, 2014. 130(25): p. e344–426.

37. Surma, M.A., et al., An automated shotgun lipidomics platform for high throughput, comprehensive, and quantitative analysis of blood plasma intact lipids. Eur J Lipid Sci Technol, 2015. 117(10): p. 1540–1549.

38. Aimo, L., et al., The SwissLipids knowledgebase for lipid biology. Bioinformatics, 2015. 31(17): p. 2860–6.

39. Herzog, R., et al., LipidXplorer: a software for consensual cross-platform lipidomics. PLoS One, 2012. 7(1): p. e29851.

40. Herzog, R., et al., A novel informatics concept for high-throughput shotgun lipidomics based on the molecular fragmentation query language. Genome Biol, 2011. 12(1): p. R8.

41. Bolstad, B.M., et al., A comparison of normalization methods for high density oligonucleotide array data based on variance and bias. Bioinformatics, 2003. 19(2): p. 185–93.

42. Petri, M., et al., Derivation and validation of the Systemic Lupus International Collaborating Clinics classification criteria for systemic lupus erythematosus. Arthritis Rheum, 2012. 64(8): p. 2677–86.

43. Benjamini, Y. and Y. Hochberg, Controlling the False Discovery Rate - a Practical and Powerful Approach to Multiple Testing. Journal of the Royal Statistical Society Series B-Statistical Methodology, 1995. 57(1): p. 289–300.

44. Kuhn, M., Building Predictive Models in R Using the caret Package. Journal of Statistical Software, 2008. 28(5): p. 1–26.

45. Robin, X., et al., pROC: an open-source package for R and S plus to analyze and compare ROC curves. BMC Bioinformatics, 2011. 12.

46. Burnett, J.R., A.J. Hooper, and R.A. Hegele, Remnant Cholesterol and Atherosclerotic Cardiovascular Disease Risk. J Am Coll Cardiol, 2020. 76(23): p. 2736–2739.

47. Petri, M.A., E. Barr, and L.S. Magder, Development of a systemic lupus erythematosus cardiovascular risk equation. Lupus Sci Med, 2019. 6(1): p. e000346.

48. McMahon, M., B.H. Hahn, and B.J. Skaggs, Systemic lupus erythematosus and cardiovascular disease: prediction and potential for therapeutic intervention. Expert Rev Clin Immunol, 2011. 7(2): p. 227–41.

49. Hak, A.E., et al., Systemic lupus erythematosus and the risk of cardiovascular disease: results from the nurses’ health study. Arthritis Rheum, 2009. 61(10): p. 1396–402.

50. Zeller, C.B. and S. Appenzeller, Cardiovascular disease in systemic lupus erythematosus: the role of traditional and lupus related risk factors. Curr Cardiol Rev, 2008. 4(2): p. 116–22.

